# New and Increasing Rates of Adverse Events Can be Found in Unstructured Text in Electronic Health Records using the Shakespeare Method

**DOI:** 10.1101/2021.01.12.21249674

**Authors:** Roselie A. Bright, Katherine Dowdy, Summer K. Rankin, Sergey V. Blok, Lee Anne Palmer, Susan J. Bright-Ponte

## Abstract

**Background:** Text in electronic health records (EHRs) and big data tools offer the opportunity for surveillance of adverse events (patient harm associated with medical care) (AEs) in the unstructured notes. Writers may explicitly state an apparent association between treatment and adverse outcome (“attributed”) or state the simple treatment and outcome without an association (“unattributed”). We chose to study EHRs from 2006-2008 because of known heparin contamination during this timeframe. We hypothesized that the prevalence of adulterated heparin may have been widespread enough to manifest in EHRs through symptoms related to heparin adverse events, independent of clinicians’ documentation of attributed AEs.

**Objective:** Use the Shakespeare Method, a new unsupervised set of tools, to identify attributed and unattributed potential AEs using the unstructured text of EHRs.

**Methods:** We studied 21,287 adult critical care admissions divided into three time periods. Comparisons of period 3 (7/2007 to 6/2008) to period 2 (7/2006 to 6/2007) were used to find admissions notes to review for new or increased clinical events by generating Latent Dirichlet Allocation topics among words in period 3 that were distinct from period 2. These results were further explored with frequency analyses of periods 1 (7/2001 to 6/2006) through 3.

**Results:** Topics represented unattributed heparin AEs, other medical AEs, rare medical diagnoses, and other clinical events; all were verified with EHRs notes review and frequency analysis. The heparin AEs were not attributed in the notes, diagnosis codes, or procedure codes. Somewhat different from our hypothesis, heparin AEs increased in prevalence from 2001 through 2007, and decreased starting in 2008 (when heparin AEs were being published).

**Conclusions:** The Shakespeare Method could be a useful supplement to AE reporting and surveillance of structured EHRs data. Future improvements should include automation of the manual review process.

## INTRODUCTION

Avoidable patient harm continues to be a significant problem [1]. To learn of patient harm known as adverse events (AEs) related to products it regulates, FDA relies on spontaneous reports from manufacturers, healthcare providers, and the general public. Published deficiencies of these reports [2-10] include well known biases in reporting. Now that electronic healthcare records (EHRs) are very common [11] and seen as more informative than billing codes from payment claims [6, 12, 13] we have an opportunity to leverage them for automated surveillance of AEs [6, 14, 15].

Many methods for finding AEs in text [6, 7, 9, 16-38] rely on predefining the possible AEs. Writers may explicitly state an apparent association between treatment and adverse outcome (“attributed”) or state the simple treatment and outcome without an association (“unattributed”). More critically, attributed and unattributed potential AEs (PAEs) may not necessarily be captured in structured data (e.g., diagnosis and procedure codes) [14, 23, 39].

Many medical care AEs occur at higher frequency in hospital critical care settings, related to complex illnesses, invasive procedures, and relatively long lists of treatments [40, 41]. In previous work, we performed a comparison of transfused to non-transfused admissions to critical care at a major teaching hospital [42] that successfully found potential blood transfusion adverse events, while addressing many published challenges (such as synonyms, overlapping meanings, and nonstandard terms) with using unstructured EHRs text [5, 11, 14, 19, 23].

We hoped the Shakespeare Method [42] would overcome the challenges of EHRs text to detect not only clinical and administrative changes but also trending potential AEs (PAEs), including heparin contamination PAEs which were first reported early in 2008 [43].

## METHODS

The Shakespeare strategy is to find unusual, significant words that were new or increased in the most recent time period, use topic analysis to find words that tended to occur together, examine admissions that were prominent for topics of interest, and then evaluate how well the topics performed [42].

### Study Population

We used EHRs for critical care admissions within an adult hospital, Beth Israel Deaconess Medical Center, Boston, MA the Medical Information Mart for Intensive Care III (MIMIC-III) [35, 44], which used one medical record system in 2001-2008 and another afterwards. We received the real dates, within several weeks, for the earlier data. MIMIC III is publicly available to those meeting human subjects research requirements. The research was designated not human subjects research by the FDA Institutional Review Board under the Code of Federal Regulations [45].

We wanted to simulate real-time analysis to find new or increasing events in the most recent time period. MIMIC-III data were collected with two sequential EHRs, so we selected the longer, earlier period of exclusive use of the earlier EHRs (7/2001-6/2008). We restricted the admissions to patients > 16 years old because this was a hospital for adults.

### Preprocessing

We concatenated in chronological order all text notes for a hospital admission into a document. We removed the personally identifying information mask string and lowercased the text, and retained punctuation, numerals and stop words (because they convey clinical information and are sometimes components of abbreviations).

Since our methods would be based on the frequencies of words, we eliminated duplicate sentences because they do not represent additional information and give weight to variable personal duplication practices. We removed widespread duplicated sentences and lists within the notes, using Bloatectomy [46].

### Word Extraction

We utilized sci-kit learn’s CountVectorizer [47, 48] to convert each document into a bag of words vector where each dimension is represented by the frequency of each n-gram present in the document (see Figure 1a and 1b). Details are in Table 1.

**Table 1.**
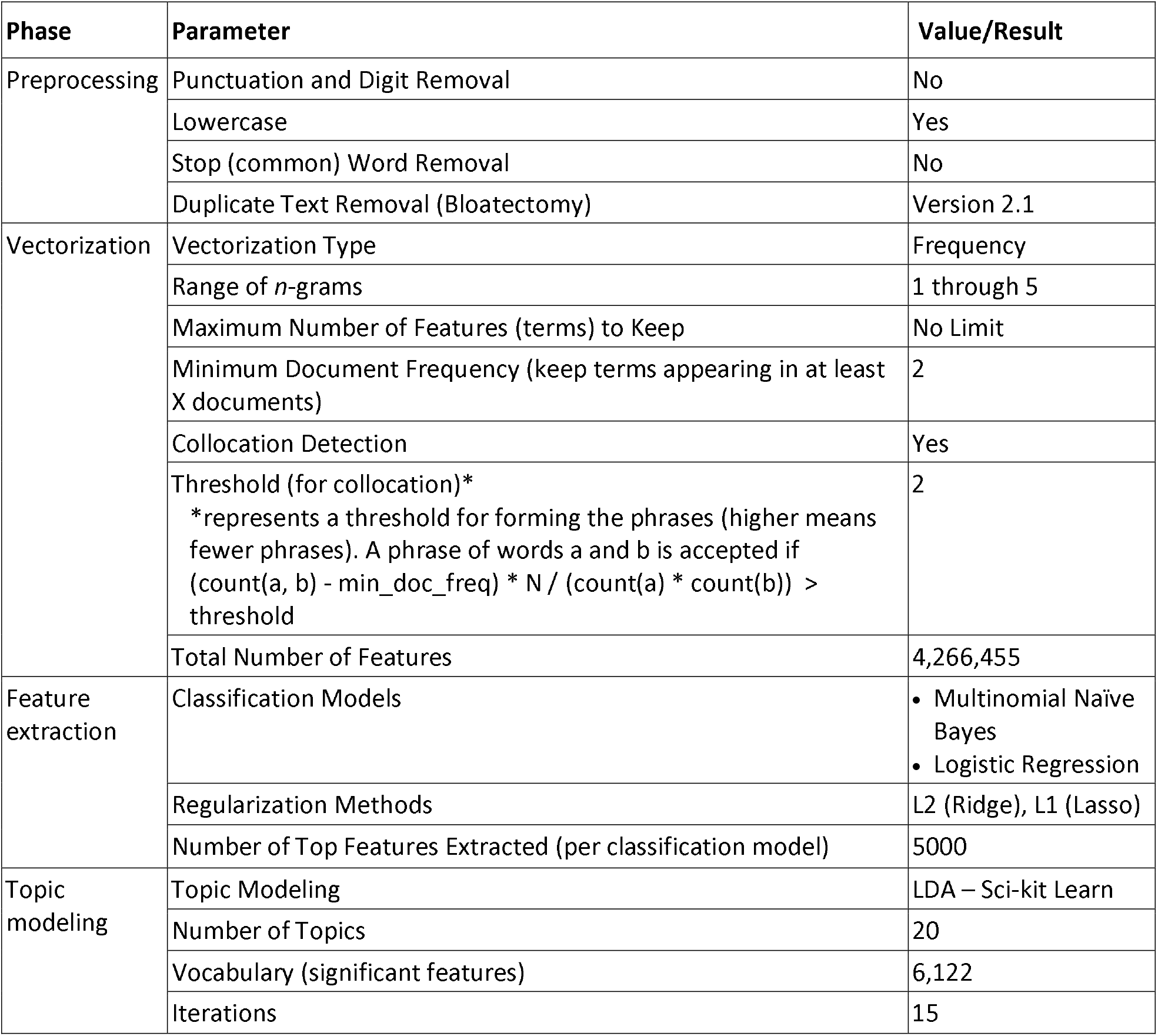
Preprocessing/Model Parameters.

| Phase | Parameter | Value/Result |
| --- | --- | --- |
| Preprocessing | Punctuation and Digit Removal | No |
|  | Lowercase | Yes |
|  | Stop (common) Word Removal | No |
|  | Duplicate Text Removal (Bloatectomy) | Version 2.1 |
| Vectorization | Vectorization Type | Frequency |
|  | Range of *n*-grams | 1 through 5 |
|  | Maximum Number of Features (terms) to Keep | No Limit |
|  | Minimum Document Frequency (keep terms appearing in at least X documents) | 2 |
|  | Collocation Detection | Yes |
| | Threshold (for collocation)*<br>*represents a threshold for forming the phrases (higher means fewer phrases). A phrase of words a and b is accepted if $(\text{count}(a, b) - \text{min\_doc\_freq}) * N / (\text{count}(a) * \text{count}(b)) > \text{threshold}$ | 2 |
|  | Total Number of Features | 4,266,455 |
| Feature extraction | Classification Models | • Multinomial Naïve Bayes<br>• Logistic Regression |
|  | Regularization Methods | L2 (Ridge), L1 (Lasso) |
|  | Number of Top Features Extracted (per classification model) | 5000 |
| Topic modeling | Topic Modeling | LDA – Sci-kit Learn |
|  | Number of Topics | 20 |
|  | Vocabulary (significant features) | 6,122 |
|  | Iterations | 15 |

**Figure 1.**
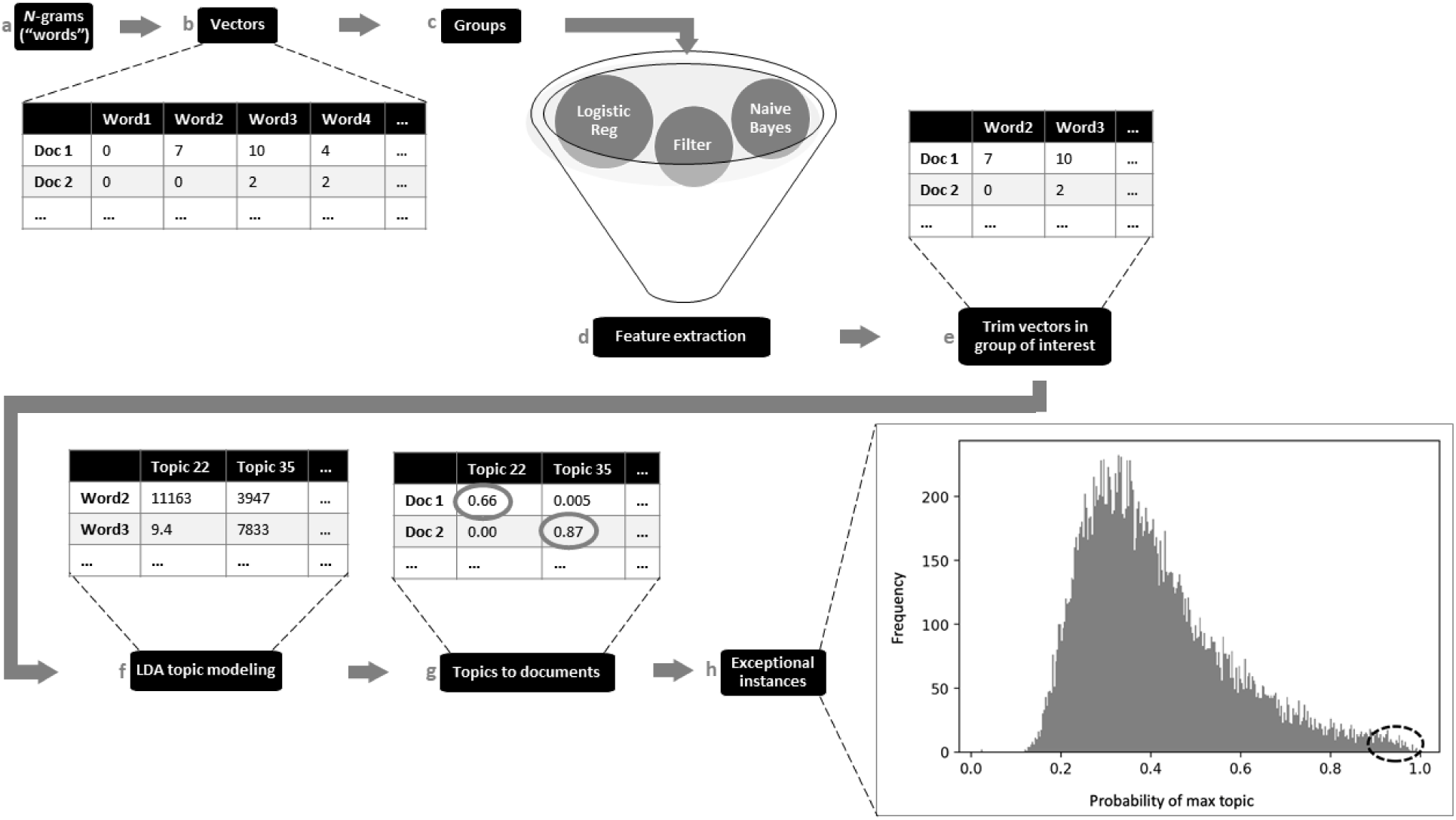
Word selection and topic modeling process with truncated examples.

We then divided the study population into three cohorts: (Period 1) admissions starting between 7/1/2001 and 6/30/2006 (14,410 documents); (Period 2) 7/1/2006-6/30/2007 (3,581 documents), and (Period 3) 7/1/2007-6/30/2008 (3,296 documents).

To focus on new or increasing AEs, we reduced the number of words to analyze by filtering by whether they were unusual and increasing (or new) in period 3 compared to period 2 (see Figures 1c, 1d and 2a). We adopted two parallel approaches, shown in Figure 2: through binary classification of the notes, and analysis of term frequency between periods 3 and 2.

**Figure 2.**
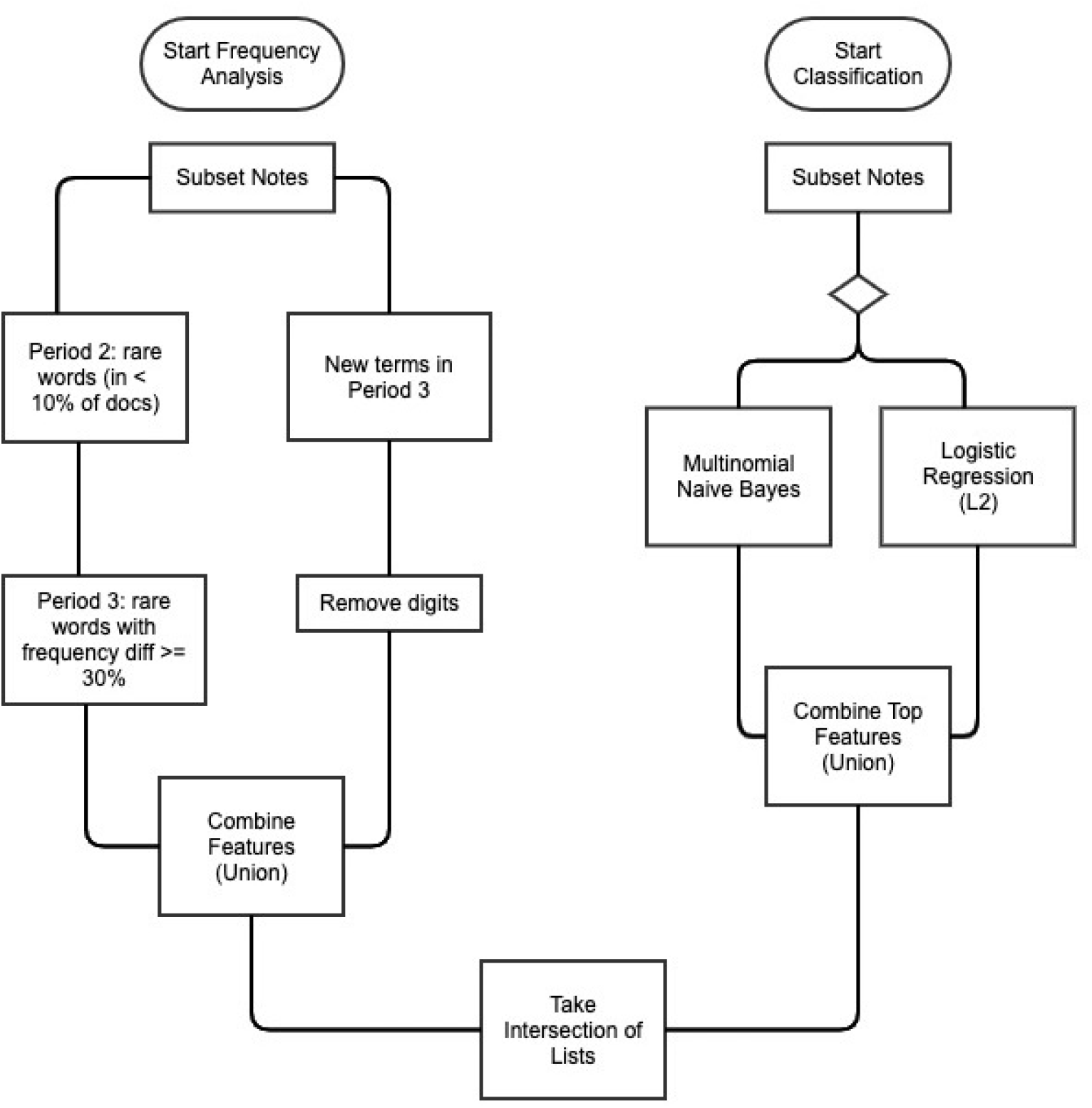
Feature extraction flowchart. This demonstrates the two parallel processes for extracting relevant features prior to topic modeling on the notes: term frequency analysis and binary classification of notes.

For the binary classification, we fit two classification models: logistic regression (LR) with L2 / ridge regularization [49] and multinomial naïve Bayes (NB) [50, 51]. Model evaluation found LR outperformed NB (with a weighted average F1 score of 0.76 compared to NB’s weighted average F1 of 0.69), but that NB more effectively identified completely new terms in the target time period.

After evaluating the models, we re-fit both models without a train-test split on the entire 24-month dataset and combined the top 5,000 features from LR (those with the highest positive coefficient, associated with the positive target class) and the top 5,000 features from NB (those with the lowest log probability ratio). Combining lists resulted in a set of 9,896 terms.

We used frequency analysis to find emerging rare clinical events. We identified two groups of terms: those which appeared in fewer than 10% of documents in period 2 and saw a 30% increase in raw frequency in period 3, and any terms that never appeared in period 2 and did appear in period 3. For those new terms appearing in period 3, we filtered out digit-only terms (a large number of terms in this group).

For the final feature set, we took the intersection of terms identified from the binary classification and frequency analysis processes. This resulted in 6,122 significant terms identified from the initial 117,049 unique terms in documents from period 3 (5.2% of terms). We re-vectorized (Figure 1e) the 12-month corpus from period 3 using the combined feature list as our vocabulary (which has the effect of filtering the notes to only include terms in the vocabulary).

### Topic Modeling and Interpretation

The co-occurrence of words in documents in the last time period was analyzed with Latent Dirichlet Allocation (LDA) topic analysis [52]. We chose the final number of topics (20) based on a balance of large and small topics and at least one topic with no substantive words. We used the words with the highest scores of their relationship to topics (Figure 1f), as well as the topic document scores that indicate the probability of the topic fit for a document (Figure 1g), to explore topic meanings. We manually read the three top-scoring documents for each topic (Figure 1h).

### Statistical Analysis of Words and Codes Suggested by Manual Review of the Topics

Documents from selected individual admissions, as well as summary data from 7/2001 to 6/2008 were used to evaluate whether any topics formed around AEs. Most topics inspired time plots of selected words, diagnosis codes, or procedure codes through periods 1, 2, and 3. Slopes were analyzed for changes [53, 54].

For this report, out of concern for patient privacy we substituted generic words (such as “condition01”, “condition02”, etc.) for rare conditions, drugs, events, and languages because the year of admission is being presented. Related substitute words (e.g., “condition09a”, “condition09b”) were used for synonyms.

## RESULTS

Table 2 shows the statistics for each topic. The strength of the maximum word score in a topic roughly corresponded with the number of admissions that had strong matches with the topic. The words in many of the topics seem to readily suggest interpretations, for example: long complex stay (topic 18), heart problem (3), trauma (19), cardiac catheterization (7), brain (1), cardiac catheterization (17), abdomen (12), uterus (16), and a foreign language (2). The other topics seemed broad.

**Table 2.** Topics sorted by the maximum word score in the topic, with the top 20 substantive words, the maximum topic match score among admissions, and distribution of the topic match scores among admissions. “Substantive” words had topic scores above the minimum topic score. “Max” means maximum.

| Topic |  |  | Max topic match score among admissions | # Admissions in topic score range |  |  |  |  |
| --- | --- | --- | --- | --- | --- | --- | --- | --- |
| Topic # | Top 20 substantive terms | Max word score |  | ≥0.03 | ≥0.50 | ≥0.2 to <0.5 | ≥0.1 to <0.2 | ≥0.03 to <0.1 |
| 18 | for, hr, plan, vent, intubated, cont, today, skin, are, family, per, support, increased, off, goal, iv, placed, trach, foley, pain | 75372 | 0.99 | 1793 | 505 | 623 | 326 | 339 |
| 3 | for, hr, pain, bp, are, you, iv, family, time, ccu, per, sats, note, heart, micu, received, skin, if, acute, plan | 42070 | 1.0 | 2224 | 912 | 697 | 328 | 287 |
| 19 | for, are, pain, you, comparison, acute, upper, evaluate, iv, trauma, hospital, if, note, time, large, level, pleural, wbc, read, throughout | 39731 | 1.0 | 2089 | 355 | 880 | 468 | 386 |
| 7 | for, are, pain, pleural, cabg, hr, plan, per, comparison, off, bp, pericardial, time, neo, iv, heart, md, mm, mr, catheter | 30722 | 1.0 | 1686 | 589 | 321 | 319 | 457 |
| 1 | for, are, family, subarachnoid, mm, comparison, pain, iv, occipital, sdh, large, evaluate, plan, cont, acute, craniotomy, per, hr, note, goal | 12352 | 1.0 | 749 | 181 | 235 | 118 | 215 |
| 4 | catheter, pleural, for, pain, jp, [pain-reliever], placed, large, into, pigtail, hr, cont, french, increased, are, pseudoaneurysm, upper, skin, iv, comparison | 3523 | 0.54 | 683 | 1 | 75 | 180 | 427 |
| 17 | for, are, mca, into, time, catheter, arteriogram, occlusion, mm, acute, french, ica, iv, placed, territory, large, cont, comparison, goal, family | 3462 | 0.77 | 534 | 39 | 99 | 127 | 269 |
| 12 | [condition01], section, gynecology, [condition02], dystrophy, cesarean, [anti-thyroid], transabdominal, [event01], lmp, wk, [procedure01], [progesterone], prenatal, [condition03], [condition04], [antispasmodic], enteropathy, [condition05], [condition06] | 216 | 0.22 | 31 | 0 | 1 | 7 | 23 |
| 11 | pentobarb, pentobarbital, cmv, encasement, prison, [condition07], satellite, hematologic, rent, [condition08], [condition09a], [condition09b], [antibiotic], federal, bleach, [device01], allergic, [rare-word01], cluster, [rare-word02] | 75 | 0.11 | 26 | 0 | 0 | 1 | 25 |
| 5 | [rare words, misspelled words] | 63 | 0.05 | 1 | 0 | 0 | 0 | 1 |
| 15 | [rare words, misspelled words] | 36 | 0.13 | 2 | 0 | 0 | 2 | 0 |
| 16 | [rare words, misspelled words] | 15 | 0.11 | 2 | 0 | 0 | 1 | 1 |
| 6 | [rare words, misspelled words] | 14 | 0.02 | 0 | 0 | 0 | 0 | 0 |
| 10 | [rare words, misspelled words] | 11 | 0.06 | 2 | 0 | 0 | 0 | 2 |
| 0 | [rare word] | 10 | 0.04 | 1 | 0 | 0 | 0 | 1 |
| 2 | [rare words, foreign language words, misspelled words] | 9 | 0.12 | 3 | 0 | 0 | 1 | 2 |
| 14 | [rare words, misspelled words] | 8 | 0.03 | 1 | 0 | 0 | 0 | 1 |
| Topic # | Top 20 substantive terms | Max word score | | $\geq 0.03$ | $\geq 0.50$ | $\geq 0.2$ to $< 0.5$ | $\geq 0.1$ to $< 0.2$ | $\geq 0.03$ to $< 0.1$ |
| 9 | [rare words, misspelled words] | 7 | 0.07 | 2 | 0 | 0 | 0 | 2 |
| 13 | [rare words, misspelled words] | 6 | 0.06 | 3 | 0 | 0 | 0 | 3 |
| 8 |  | 0 | 0.00 | 0 | 0 | 0 | 0 | 0 |

### Common topics

For the most common topics, the admissions with the top three topic match scores are summarized in Table 3. For the topics with words that suggested an interpretation, the records supported the interpretations. For the other topics, the records suggested interpretations that were consistent with the top words. Each of the three top scoring admissions within a topic were quite similar to each other (an indication that the topics were coherent and the model was working correctly; with the exception of the third admission in topic 3).

**Table 3.**
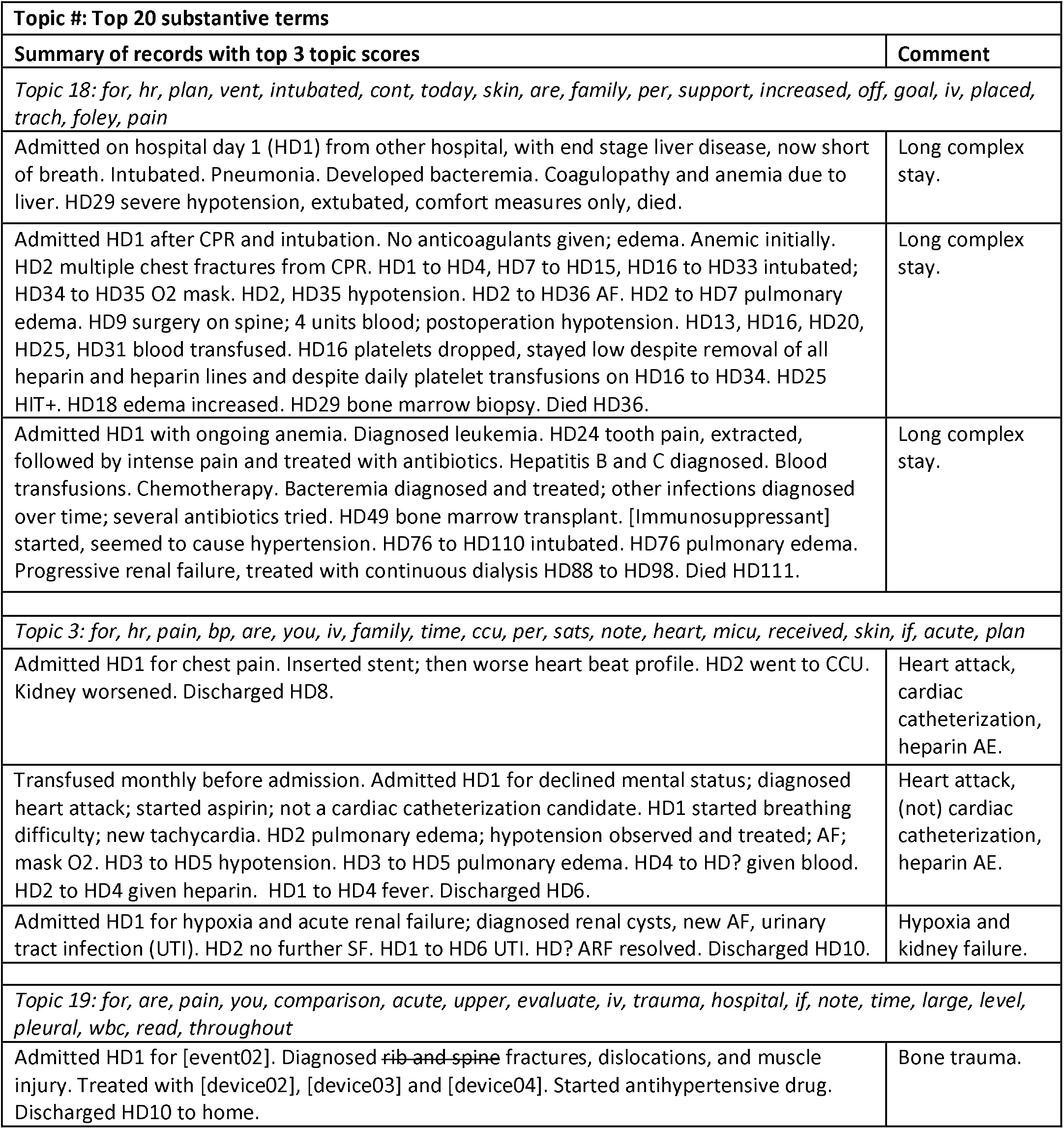
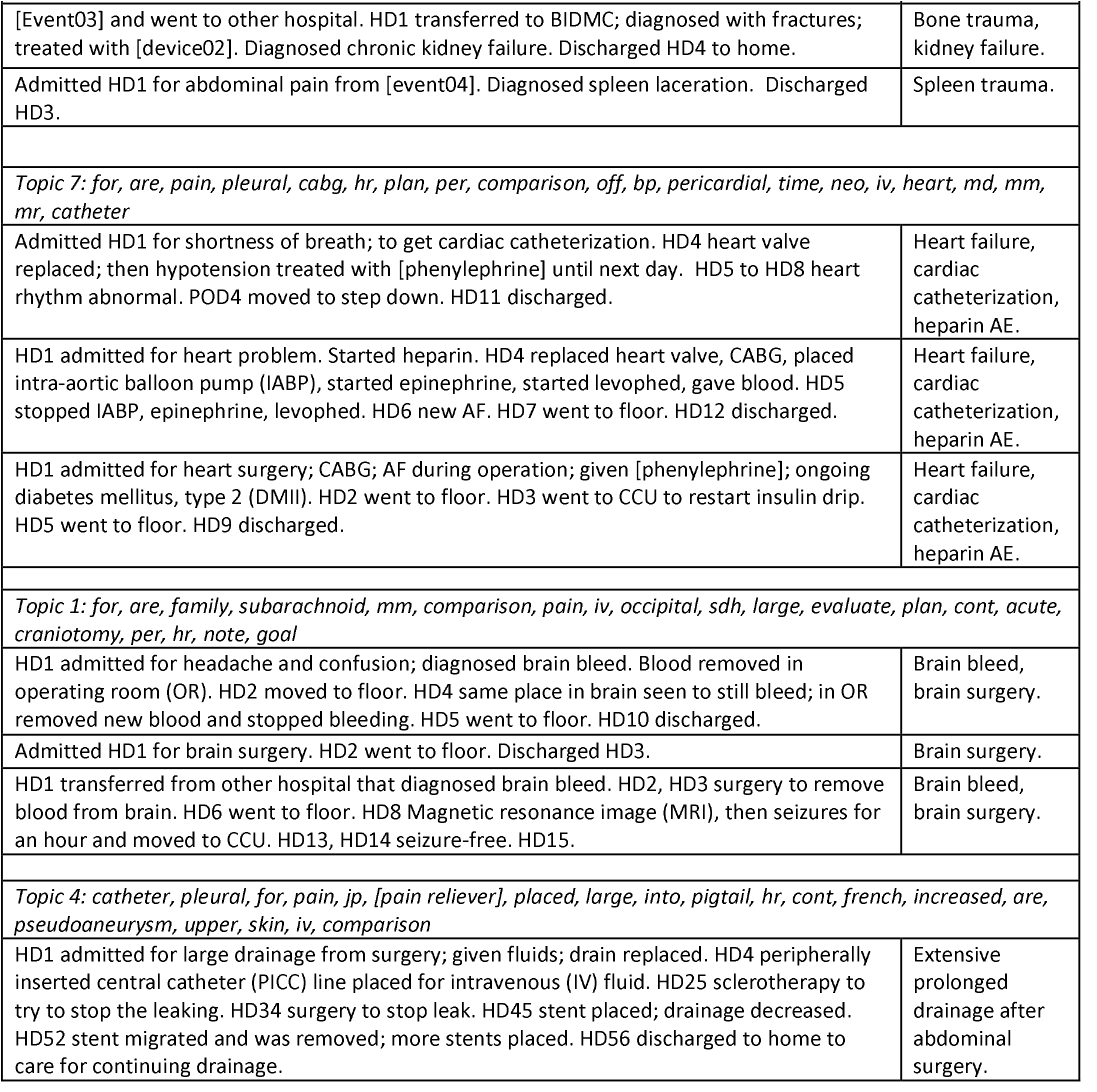
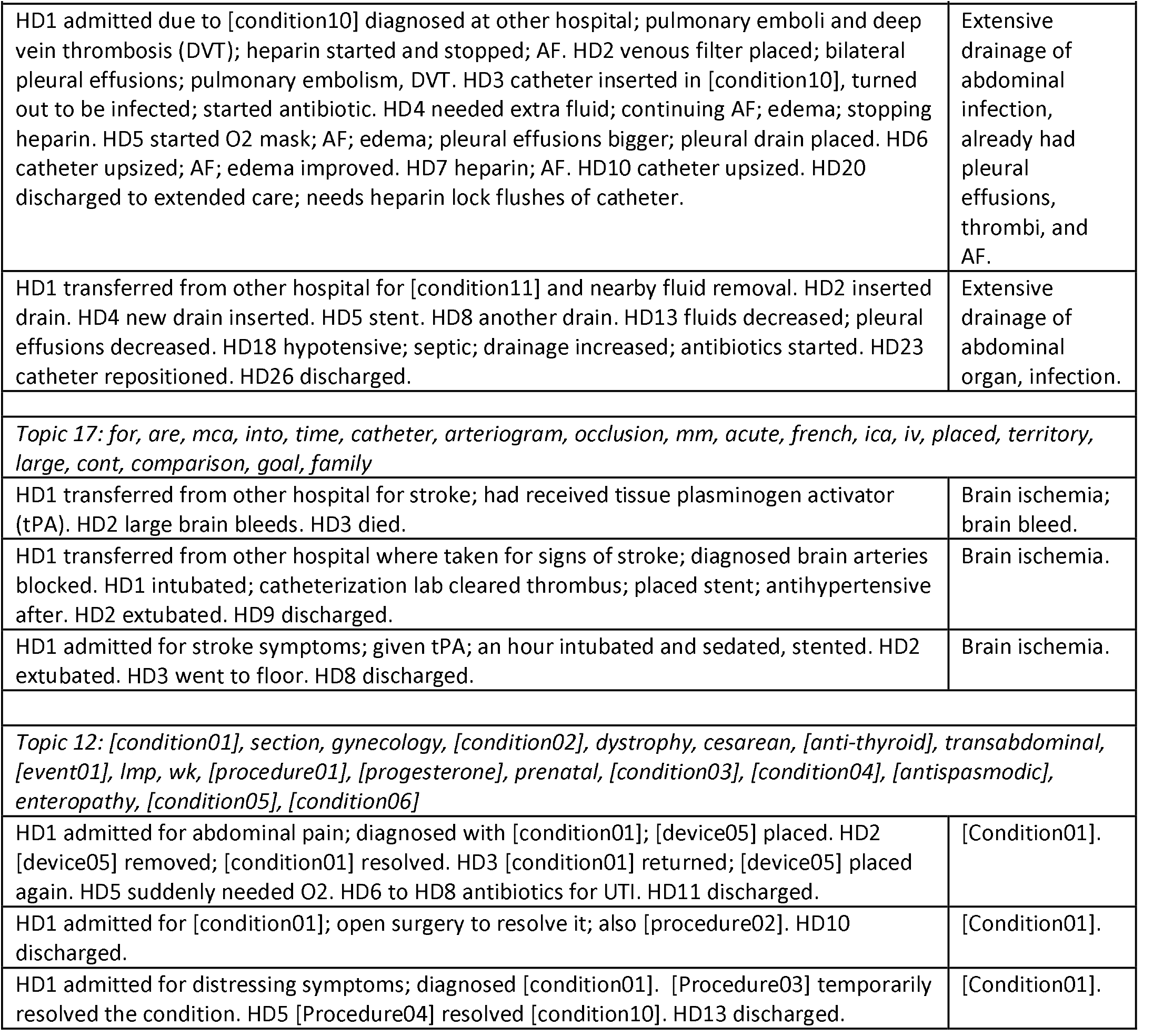
Summaries of the admissions with the top three topic match scores, for the most common topics. “HD” is hospital day. “Intubated” and “extubated” refer to starting and ending mechanical ventilation.

The top three scoring documents for topic 18 described long complex stays, which included large numbers of notes. The general words in the topic (“for”, “hr”, “plan”, “cont”, “today”, “skin”, and “are”) are nearly ubiquitous in periods 2 and 3. The words indicating mechanical ventilation (“vent”, “intubated”, and “trach”) were present in between 51% and 58% of the admissions per quarter in periods 2 and 3, with a slight, not clinically significant increase for period 3. The lengths of stay and numbers of notes also did not vary between periods 2 and 3.

We noticed that among the five records in Table 3 that mentioned cardiac catheterization, all mentioned explicit or implied dosing with heparin followed the same day with hypotension that required treatment (heparin is generally involved with cardiovascular procedures) [55].

Topics 3 and 7 both have cardiac catheterization for heart problems in common; for five out of six instances, the procedure or heparin administration was followed by hypotension (four instances) that needed to be treated or heart rhythm deterioration (one instance). To investigate whether these potential heparin AEs were increasing 7/2001-6/2008, we plotted two measures of exposure (invasive cardiac procedure code and “heparin”) and a measure of AE (“hypotension”). The proportion of admissions that had invasive cardiovascular procedure codes (see Figures 3a and b) declined overall (see Figure 3a), but had a local increase in period 3, compared to period 2. In contrast to the procedures, the words “heparin” and “hypotension” showed an overall rough increase over the entire timeframe. We also noticed that the proportion of admissions with invasive cardiology codes that had the word “hypotension” increased gradually over time (Figures 3a and b), followed by a drop in the last quarter; the pattern was similar and weaker for the proportion of admissions with “heparin” that also had “hypotension”. There was a decrease in “hypotension” in the last quarter, both as a proportion of all admissions, and as a proportion of either indicator of having been exposed to heparin.

**Figure 3.**
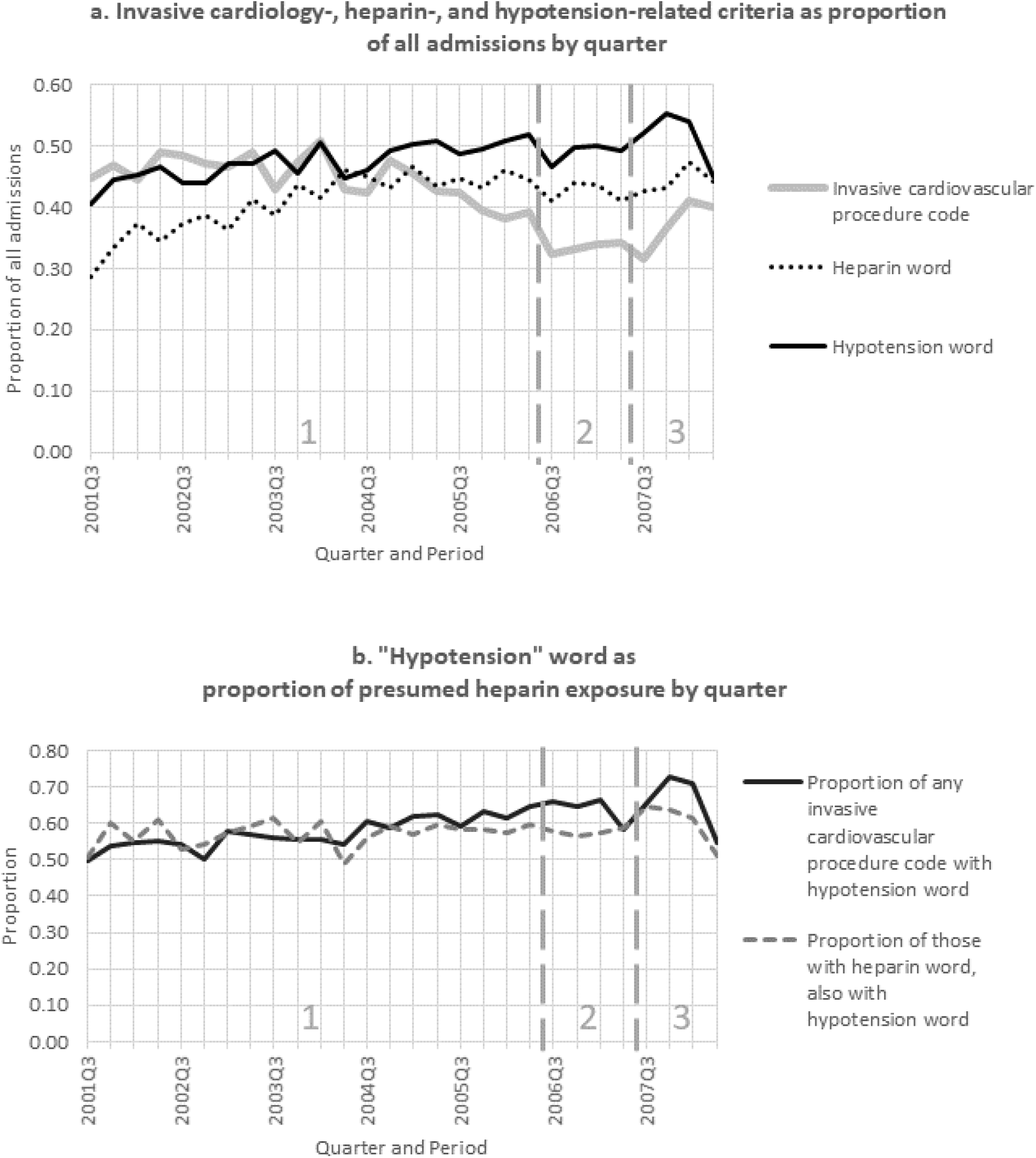
Heparin and hypotension. **Figure 3a. Invasive cardiology-, heparin-, and hypotension-related criteria as proportion of all admissions.** Invasive cardiology is presumed to involve heparin treatment. The definitions are listed in “Results eFigures 2 to 6 and figures info”. For invasive cardiovascular procedure code, slope = −0.0053 (95%CL −0.0069 to −0.0037, p<0.0001), for heparin word slope = 0.0039 (95% CI 0.0025 to 0.0054, p<0.0001), and for hypotension word slope = 0.0029 (95%CL 0.0017 to 0.0040, p<0.0001). **3b. “Hypotension” word as proportion of presumed heparin exposure.** For proportion of any invasive cardiovascular procedure code (presumed to involve heparin), slope = 0.0055 (95%CL 0.0038 to 0.0072, p<0.0001). For proportion of those with “heparin”, slope = 0.0013 (95%CL −0.00036 to 0.0030, p=0.12). Figure 3 notes: • Invasive cardiovascular code: • 3891 Arterial catheterization • 3961 Extracorporeal circulation auxiliary to open heart surgery • [3965 to 3966] • “Heparin” word • “Hypotension” word

### Other common topics

Topic 19 (and 13) corresponded with trauma. Figure 4 showed that trauma diagnosis and procedure codes increased steadily over time through periods 1-3.

**Figure 4.**
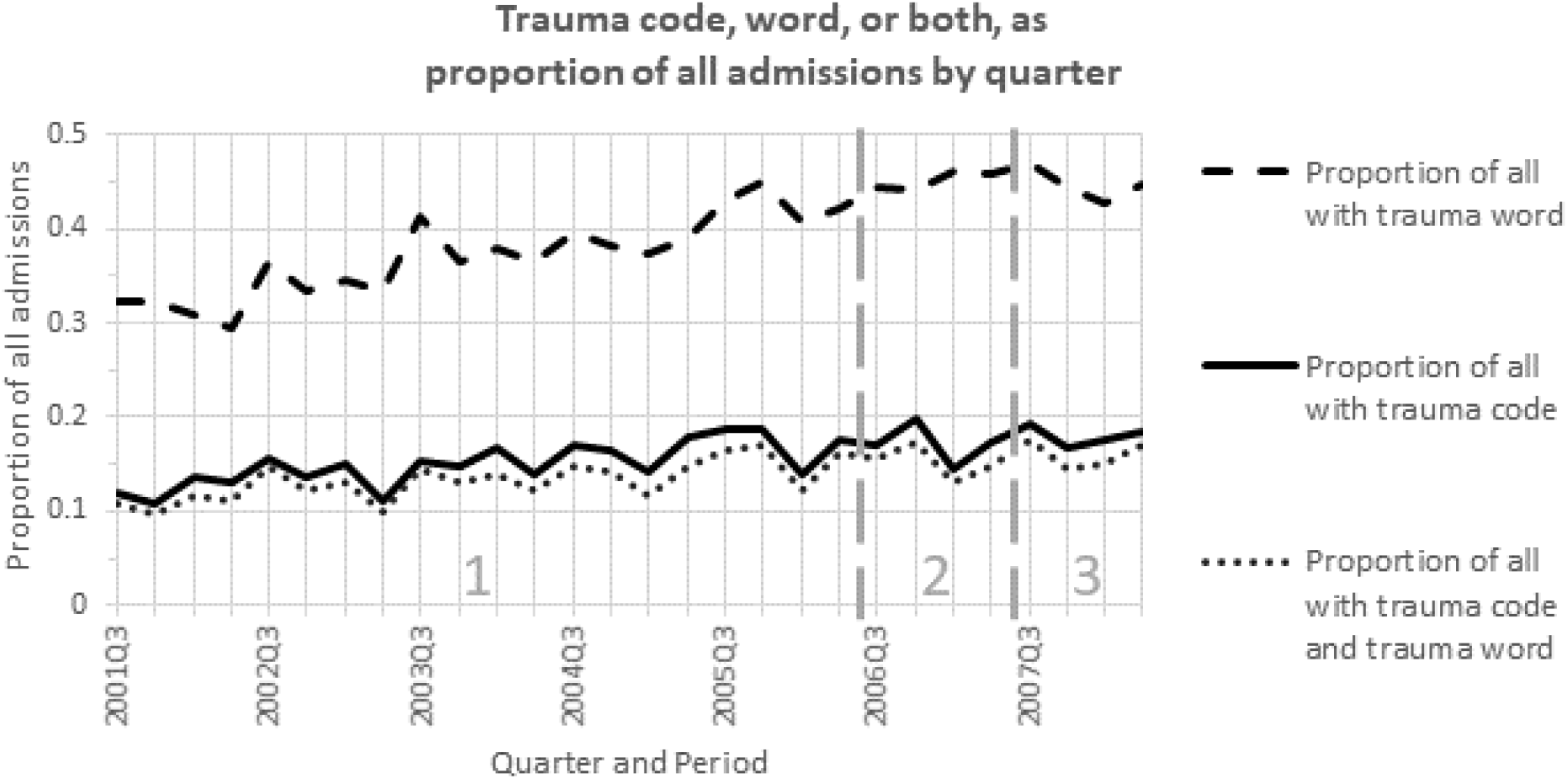
Trauma code, word, or both as proportion of all admissions by quarter. For proportion with trauma code, slope=0.0022 (95%CL 0.0014 to 0.0030), p<0.0001. For proportion with trauma word, slope=0.0057 (95%CI 0.0047 to 0.0067), p<0.0001. For proportion with both trauma code and word, slope=0.0019 (95% CL 0.0012 to 0.0027), p<0.0001. Figure 4 notes: • Trauma code, any of: • Diagnosis code 800* to 829* [Fracture] • Diagnosis code 830* to 869* [Dislocations, sprains, strains, and internal injury of cranium, chest, abdomen, pelvis] • Diagnosis code 870* to 897* [Open wound] • Diagnosis code 900* to 904* Injury to blood vessels • Diagnosis code 905* Late effects of musculoskeletal and connective tissue injuries • Diagnosis code 9060 to 9064 [Late effects of open wound, superficial injury, contusion, or crushing] • Diagnosis code 907* Late effects of injuries to the nervous system • Diagnosis code 908* Late effects of other and unspecified injuries • Diagnosis code 910* to 924* [Superficial injury and contusion with intact skin surface] • Diagnosis code 925* to 929* Crushing injury • Diagnosis code 950* to 957* Injury To Nerves And Spinal Cord • Diagnosis code 958* to 959* Certain Traumatic Complications And Unspecified Injuries • Procedure code [7670 to 7679] • Procedure code [7810 to 7819] • Procedure code [7900 to 7939] • Procedure code [7960 to 7969] • Procedure code [7990 to 7999] • Trauma word, any of: • trauma • mva • fall • mvc • contusion • fracture

The brain topic (1 and 17, combined) is centered around admissions for brain injury: either bleeding, ischemia, or trauma. Figures 5a, 5b, and 5c show that there were local increases in codes for bleeding and ischemia for period 3 compared to period 2. There were slight increases in the codes for all three types of brain injuries overall. The text words indicating these conditions showed similar trends.

**Figure 5.**
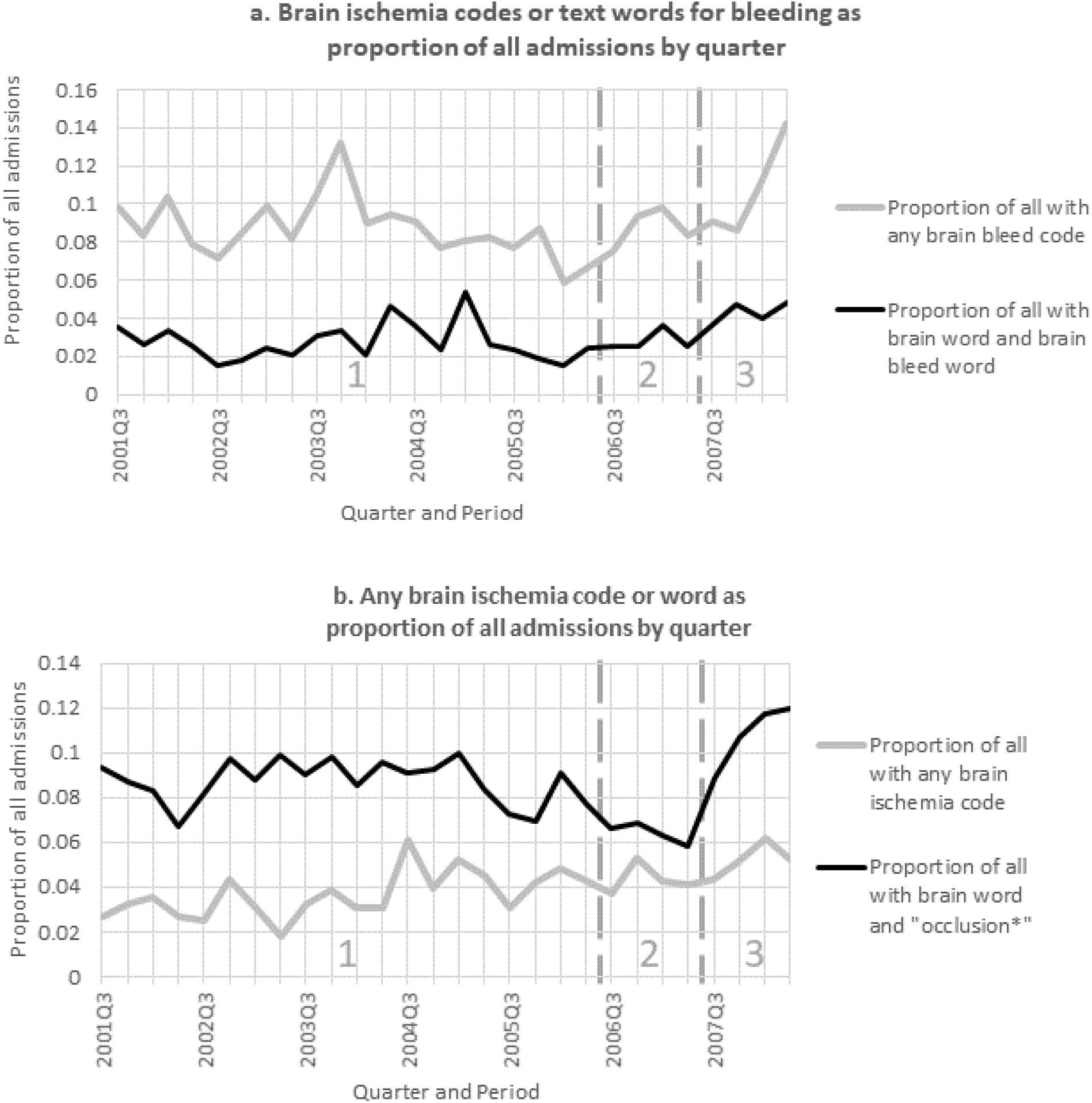
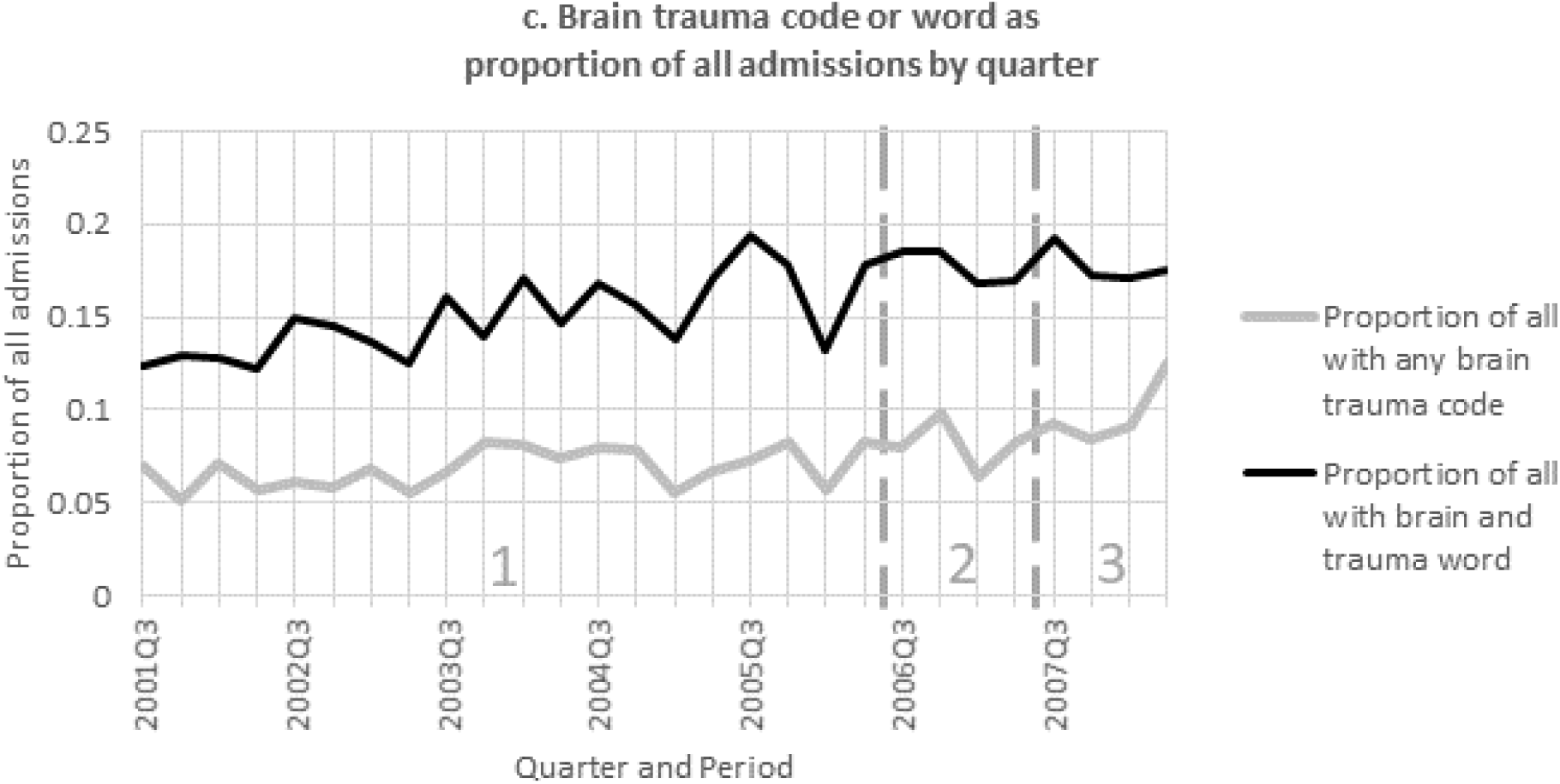
Brain ischemia codes or text words for a. bleeding, b. ischemia, and c. trauma, as proportion of all admissions by quarter. For brain bleed code, slope=0.00022 (95%CI −0.0006 to 0.0010), p=0.61. For brain word and brain bleed word, slope= 0.00039 (95%CI 0 to 0.00085), p=0.10. For brain ischemia code, slope-0.00019 (95%CI 0.00051 to 0.0013), p<0.0001. For brain word and “occlusion*”, slope = 0 (95%CI - 0.00064 to 0.00080), p=0.84. For brain trauma code, slope=0.0013 (95%CI 0.00073 to 0.0018), p<0.0001. For brain word and “trauma”, slope=0.0021 (95%CI 0.0014 to 0.0028), p<0.0001. Figure 5 notes: • Brain bleed code is any of: • Diagnosis code 3481* Anoxic brain damage • Diagnosis code 3484* Compression of brain • Diagnosis code 430* Subarachnoid hemorrhage • Diagnosis code 431 Subarachnoid hemorrhage • Diagnosis code 432 Other and unspecified intracranial hemorrhage • Diagnosis code 8042* Closed fractures involving skull or face with other bones with subarachnoid subdural and extradural hemorrhage • Diagnosis code 8043* Closed fractures involving skull or face with other bones, with other and unspecified intracranial hemorrhage • Diagnosis code 8047* Open fractures involving skull or face with other bones with subarachnoid subdural and extradural hemorrhage • Diagnosis code 8048* Open fractures involving skull or face with other bones with other and unspecified intracranial hemorrhage • Diagnosis code 852** Subarachnoid subdural and extradural hemorrhage following injury • Diagnosis code 853** Therapeutic and unspecified intracranial hemorrhage following injury • Procedure code 109 Other cranial puncture • Procedure code 110 Intracranial pressure monitoring • Procedure code 116 Intracranial oxygen monitoring • Procedure code 121 Incision and drainage of cranial sinus • Procedure code 123 Reopening of craniotomy site • Procedure code 124 Other craniotomy • Procedure code 125 Other craniectomy • Procedure code 3881 Other surgical occlusion of vessels, intracranial vessels • Brain bleed word, any of: “IPH”, “aneurysms”, or “embolize” • Brain word, any of • *occipital • *cranio* • *cepha* • mening* • *frontal • *tempero* • *pariet* • brain • *arachnoid • mca • hemiparesis • hemiplegia • Brain ischemia code is any of: • Diagnosis code 3481* Anoxic brain damage • Diagnosis code 434** Occlusion of cerebral arteries • Diagnosis code 435** Transient cerebral ischemia • Diagnosis code 4371* Other generalized ischemic cerebrovascular disease • Diagnosis code 4376* Nonpyogenic thrombosis of intracranial venous sinus • Procedure code 62 Percutaneous angioplasty of intracranial vessel(s) • Procedure code 65 Percutaneous insertion of intracranial vascular stent(s) • Procedure code 116 Intracranial oxygen monitoring • Procedure code 1754 Percutaneous atherectomy of intracranial vessel(s) • Procedure code 3811 Endarterectomy, intracranial vessels • Brain trauma code is any of: • Diagnosis code 3485* Cerebral edema • Diagnosis code 3484* Compression of brain • Diagnosis code 34939 Other dural tear • Diagnosis code 800** Fracture of vault of skull • Diagnosis code 801** Fracture of base of skull • Diagnosis code 803** Other and unqualified skull fractures • Diagnosis code 804** Multiple fractures involving skull or face with other bones • Diagnosis code 850** Concussion • Diagnosis code 851** Cerebral laceration and contusion • Diagnosis code 852** Subarachnoid subdural and extradural hemorrhage following injury • Diagnosis code 853** Other and unspecified intracranial hemorrhage following injury • Diagnosis code 854** Intracranial injury of other and unspecified nature • Procedure code 109 Other cranial puncture • Procedure code 110 Intracranial pressure monitoring • Procedure code 116 Intracranial oxygen monitoring • Procedure code 123 Reopening of craniotomy site • Procedure code 124 Other craniotomy • Procedure code 125 Other craniectomy • Procedure code 202 Other craniectomy

Topic 4 describes prolonged drainage after abdominal surgery. The index surgeries were performed before admission for two instances and during hospitalization for the third. Figure 6 shows that codes for wounds were quite infrequent. However, long patient stays with words for leaky surgical wound or catheter were more common, rose gradually over time, and had a local increase in period 3, compared to period 2.

**Figure 6.**
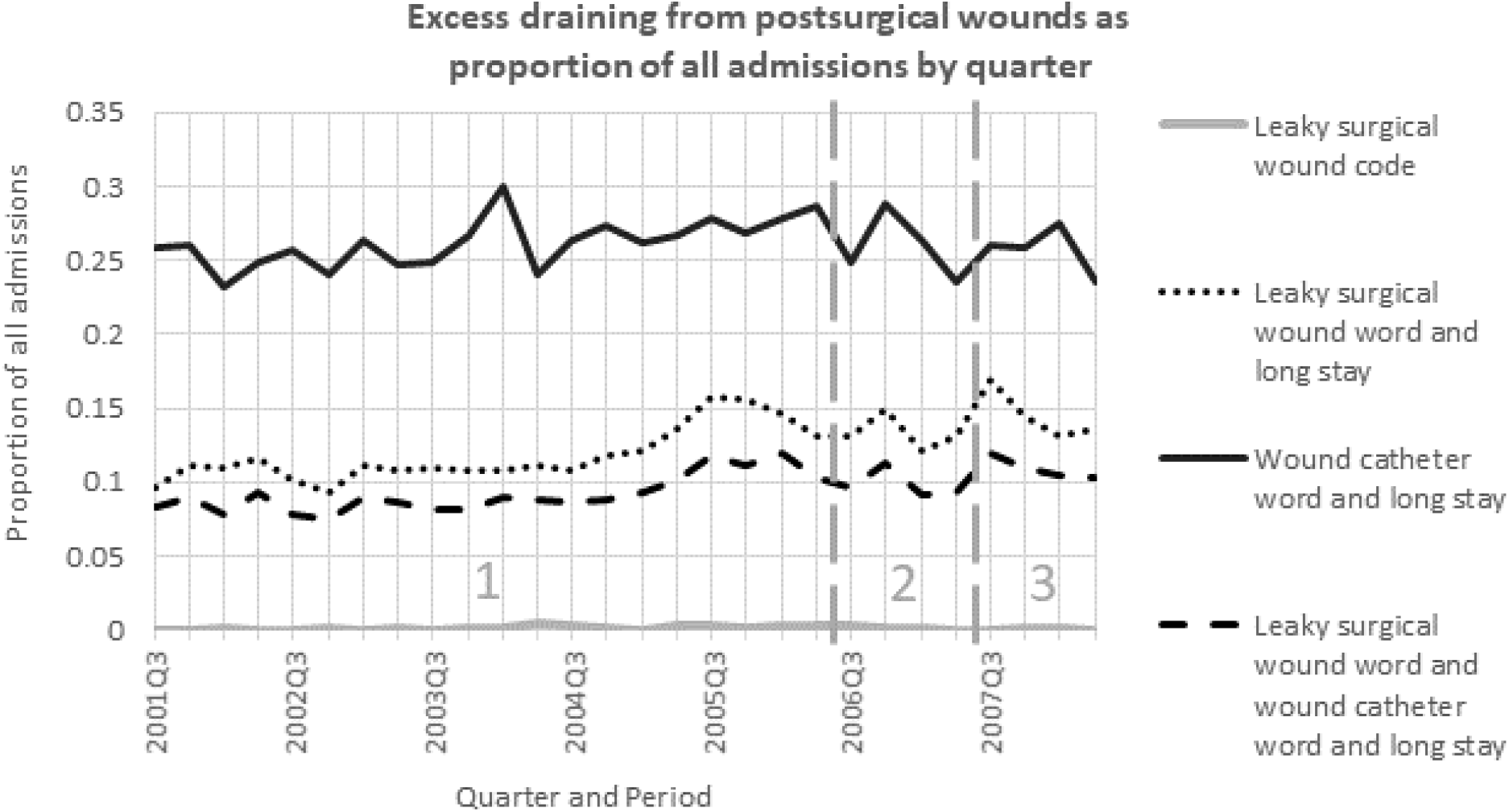
Excess draining from postsurgical wounds as proportion of all admissions by quarter. For leaky surgical wound code, slope=0.000027 (95%CI −0.000028 to 0.000082), p=0.34. For leaky surgical wound word and long stay, slope=0.0018 (95%CI 0.0012 to 0.0024), p<0.0001. For wound catheter word and long stay, slope=0.00038 (95%CI −0.00039 to 0.0012), p=0.34. For leaky surgical wound word and wound catheter word and long stay, slope=0.0011 (95%CI 0.00071 to 0.0016), p<0.0001. Figure 6eFigure 4 notes: • Leaky surgical wound word: text has “surg*” and “wound” and (“drain*” or “leak*) • Long stay: >9 days in hospital admission • Wound catheter word: “catheter”, “placed”, or “large”

Condition01 was the subject of the admissions with the top match scores for topic 12. The codes and words were generally rare for the 3 periods and showed a local increase between periods 2 and 3.

### Less common topics

Summaries of admissions with topic matching scores for the less common topics are shown in Table 4. We examined the top scoring admissions matched to topic 11 and all admissions matched to the others. All admissions in this Table had topic match scores for the index topic of <0.15 (column 2). Despite each admission in Table 4 having at least one strong topic match score for at least one of the strong topics in Table 3, the topics in Table 4 are distinct from those in Table 3. Some of the topics have admissions that have common aspects (topics 11, 10, 2, 9).

**Table 4.**
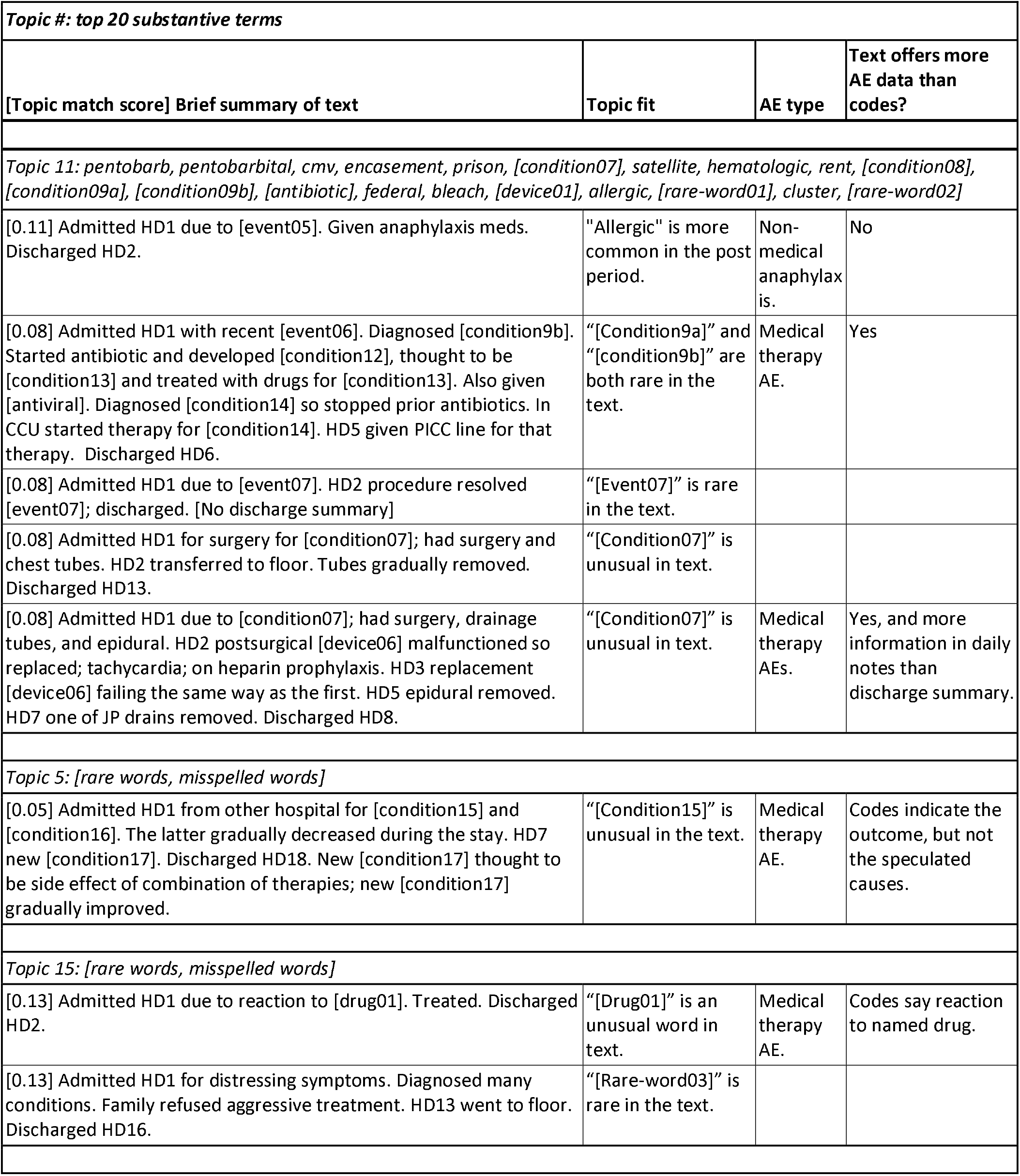
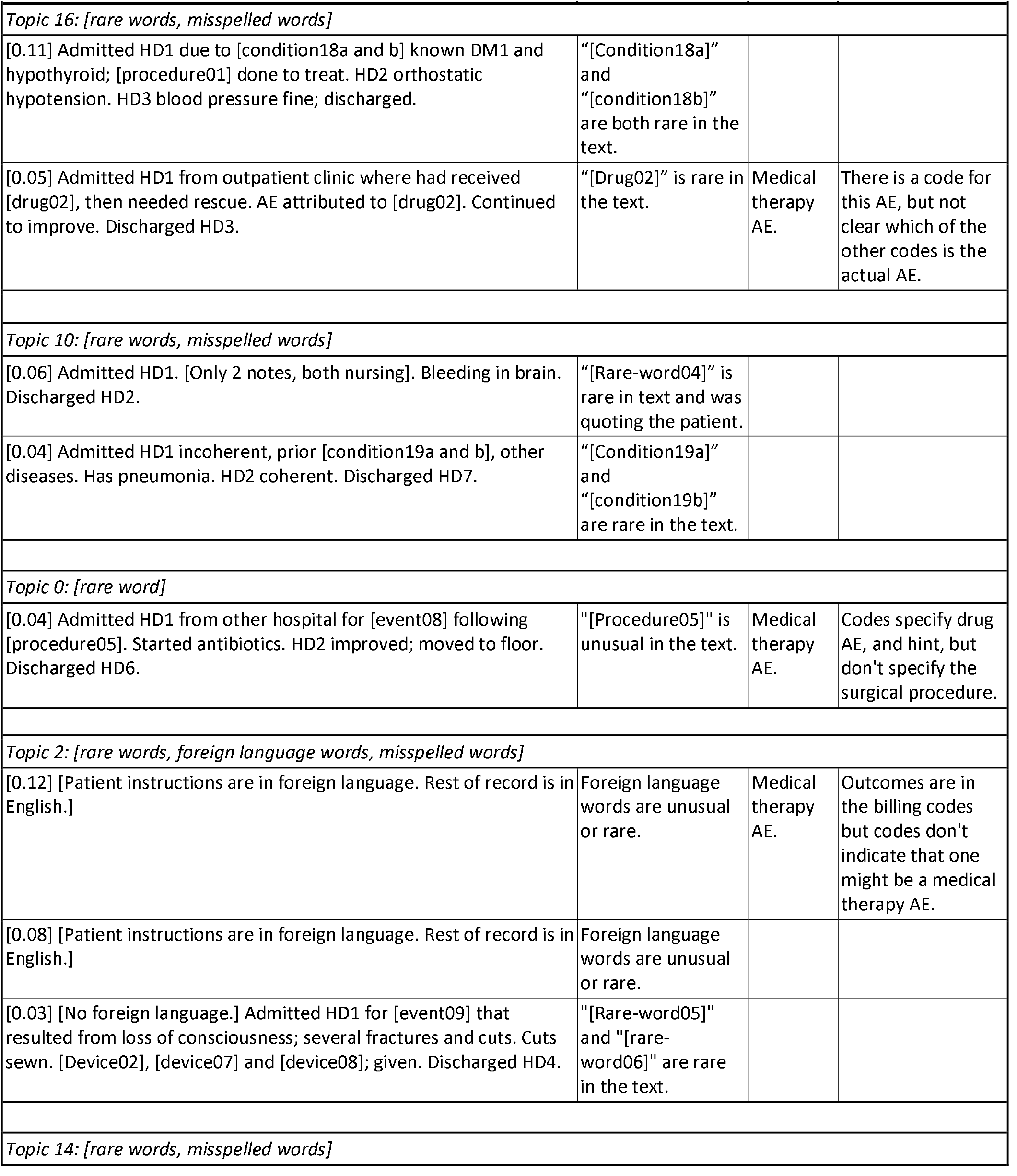
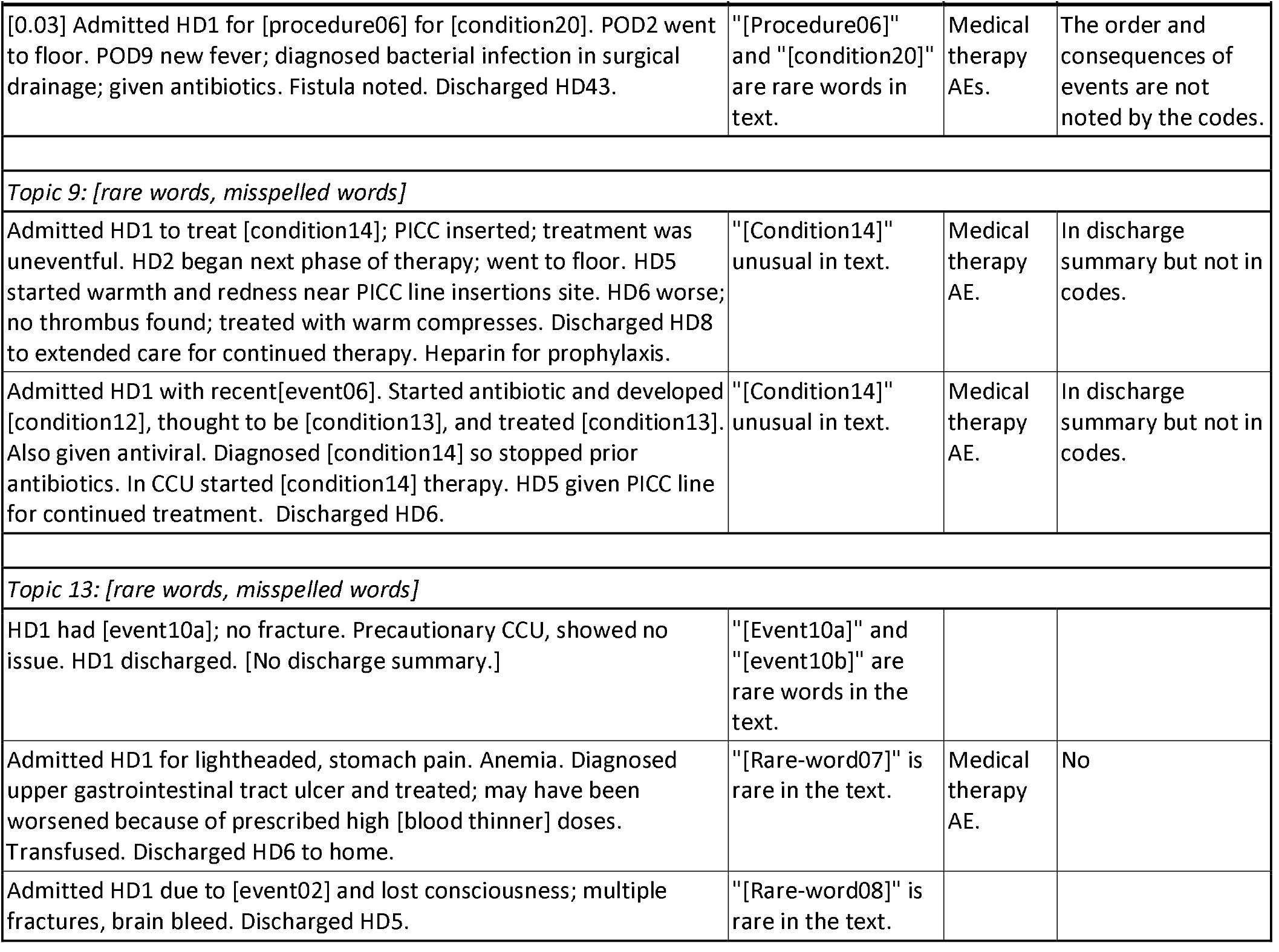
Summaries of admissions (top scoring for 20-11 and all for the other topics) with topic matching scores for the less common topics. “Unusual” means there were a few or some instances in period 1. “Rare” means there were no instances in period 1.

| **Topic #: top 20 substantive terms** |  |  |  |
| --- | --- | --- | --- |
| **[Topic match score] Brief summary of text** | **Topic fit** | **AE type** | **Text offers more AE data than codes?** |
| *Topic 11: pentobarb, pentobarbital, cmv, encasement, prison, [condition07], satellite, hematologic, rent, [condition08], [condition09a], [condition09b], [antibiotic], federal, bleach, [device01], allergic, [rare-word01], cluster, [rare-word02]* |  |  |  |
| [0.11] Admitted HD1 due to [event05]. Given anaphylaxis meds. Discharged HD2. | "Allergic" is more common in the post period. | Non-medical anaphylaxis. | No |
| [0.08] Admitted HD1 with recent [event06]. Diagnosed [condition9b]. Started antibiotic and developed [condition12], thought to be [condition13] and treated with drugs for [condition13]. Also given [antiviral]. Diagnosed [condition14] so stopped prior antibiotics. In CCU started therapy for [condition14]. HD5 given PICC line for that therapy. Discharged HD6. | "[Condition9a]" and "[condition9b]" are both rare in the text. | Medical therapy AE. | Yes |
| [0.08] Admitted HD1 due to [event07]. HD2 procedure resolved [event07]; discharged. [No discharge summary] | "[Event07]" is rare in the text. |  |  |
| [0.08] Admitted HD1 for surgery for [condition07]; had surgery and chest tubes. HD2 transferred to floor. Tubes gradually removed. Discharged HD13. | "[Condition07]" is unusual in text. |  |  |
| [0.08] Admitted HD1 due to [condition07]; had surgery, drainage tubes, and epidural. HD2 postsurgical [device06] malfunctioned so replaced; tachycardia; on heparin prophylaxis. HD3 replacement [device06] failing the same way as the first. HD5 epidural removed. HD7 one of JP drains removed. Discharged HD8. | "[Condition07]" is unusual in text. | Medical therapy AEs. | Yes, and more information in daily notes than discharge summary. |
| *Topic 5: [rare words, misspelled words]* |  |  |  |
| [0.05] Admitted HD1 from other hospital for [condition15] and [condition16]. The latter gradually decreased during the stay. HD7 new [condition17]. Discharged HD18. New [condition17] thought to be side effect of combination of therapies; new [condition17] gradually improved. | "[Condition15]" is unusual in the text. | Medical therapy AE. | Codes indicate the outcome, but not the speculated causes. |
| *Topic 15: [rare words, misspelled words]* |  |  |  |
| [0.13] Admitted HD1 due to reaction to [drug01]. Treated. Discharged HD2. | "[Drug01]" is an unusual word in text. | Medical therapy AE. | Codes say reaction to named drug. |
| [0.13] Admitted HD1 for distressing symptoms. Diagnosed many conditions. Family refused aggressive treatment. HD13 went to floor. Discharged HD16. | "[Rare-word03]" is rare in the text. |  |  |
| **[Topic match score] Brief summary of text** | **Topic fit** | **AE type** | **Text offers more AE data than codes?** |
| *Topic 16: [rare words, misspelled words]* |  |  |  |
| [0.11] Admitted HD1 due to [condition18a and b] known DM1 and hypothyroid; [procedure01] done to treat. HD2 orthostatic hypotension. HD3 blood pressure fine; discharged. | "[Condition18a]" and "[condition18b]" are both rare in the text. |  |  |
| [0.05] Admitted HD1 from outpatient clinic where had received [drug02], then needed rescue. AE attributed to [drug02]. Continued to improve. Discharged HD3. | "[Drug02]" is rare in the text. | Medical therapy AE. | There is a code for this AE, but not clear which of the other codes is the actual AE. |
| *Topic 10: [rare words, misspelled words]* |  |  |  |
| [0.06] Admitted HD1. [Only 2 notes, both nursing]. Bleeding in brain. Discharged HD2. | "[Rare-word04]" is rare in text and was quoting the patient. |  |  |
| [0.04] Admitted HD1 incoherent, prior [condition19a and b], other diseases. Has pneumonia. HD2 coherent. Discharged HD7. | "[Condition19a]" and "[condition19b]" are rare in the text. |  |  |
| *Topic 0: [rare word]* |  |  |  |
| [0.04] Admitted HD1 from other hospital for [event08] following [procedure05]. Started antibiotics. HD2 improved; moved to floor. Discharged HD6. | "[Procedure05]" is unusual in the text. | Medical therapy AE. | Codes specify drug AE, and hint, but don't specify the surgical procedure. |
| *Topic 2: [rare words, foreign language words, misspelled words]* |  |  |  |
| [0.12] [Patient instructions are in foreign language. Rest of record is in English.] | Foreign language words are unusual or rare. | Medical therapy AE. | Outcomes are in the billing codes but codes don't indicate that one might be a medical therapy AE. |
| [0.08] [Patient instructions are in foreign language. Rest of record is in English.] | Foreign language words are unusual or rare. |  |  |
| [0.03] [No foreign language.] Admitted HD1 for [event09] that resulted from loss of consciousness; several fractures and cuts. Cuts sewn. [Device02], [device07] and [device08]; given. Discharged HD4. | "[Rare-word05]" and "[rare-word06]" are rare in the text. |  |  |
| *Topic 14: [rare words, misspelled words]* |  |  |  |
| **[Topic match score] Brief summary of text** | **Topic fit** | **AE type** | **Text offers more AE data than codes?** |
| [0.03] Admitted HD1 for [procedure06] for [condition20]. POD2 went to floor. POD9 new fever; diagnosed bacterial infection in surgical drainage; given antibiotics. Fistula noted. Discharged HD43. | "[Procedure06]" and "[condition20]" are rare words in text. | Medical therapy AEs. | The order and consequences of events are not noted by the codes. |
| *Topic 9: [rare words, misspelled words]* |  |  |  |
| Admitted HD1 to treat [condition14]; PICC inserted; treatment was uneventful. HD2 began next phase of therapy; went to floor. HD5 started warmth and redness near PICC line insertions site. HD6 worse; no thrombus found; treated with warm compresses. Discharged HD8 to extended care for continued therapy. Heparin for prophylaxis. | "[Condition14]" unusual in text. | Medical therapy AE. | In discharge summary but not in codes. |
| Admitted HD1 with recent[event06]. Started antibiotic and developed [condition12], thought to be [condition13], and treated [condition13]. Also given antiviral. Diagnosed [condition14] so stopped prior antibiotics. In CCU started [condition14] therapy. HD5 given PICC line for continued treatment. Discharged HD6. | "[Condition14]" unusual in text. | Medical therapy AE. | In discharge summary but not in codes. |
| *Topic 13: [rare words, misspelled words]* |  |  |  |
| HD1 had [event10a]; no fracture. Precautionary CCU, showed no issue. HD1 discharged. [No discharge summary.] | "[Event10a]" and "[event10b]" are rare words in the text. |  |  |
| Admitted HD1 for lightheaded, stomach pain. Anemia. Diagnosed upper gastrointestinal tract ulcer and treated; may have been worsened because of prescribed high [blood thinner] doses. Transfused. Discharged HD6 to home. | "[Rare-word07]" is rare in the text. | Medical therapy AE. | No |
| Admitted HD1 due to [event02] and lost consciousness; multiple fractures, brain bleed. Discharged HD5. | "[Rare-word08]" is rare in the text. |  |  |

Fourteen PAEs evident in the notes were distributed among the less common topics: 13 related to medical therapy (6 medications, 3 medical devices, 2 procedures, and 2 combinations) and 2 non-medical. Five drug and all of the medical device PAEs are published in the product labels and/or medical literature. Nine of the PAEs occurred outside the hospital and were related to the reason for admission. The diagnosis and procedure codes generally did not give enough information to understand the specific cause and associated potential AE. Figure 7 shows that while the proportions over the 7 years of admissions with allergy and anaphylaxis words steadily decreased, the diagnosis codes for drug AEs and for surgical or procedure AEs increased slightly over time.

**Figure 7.**
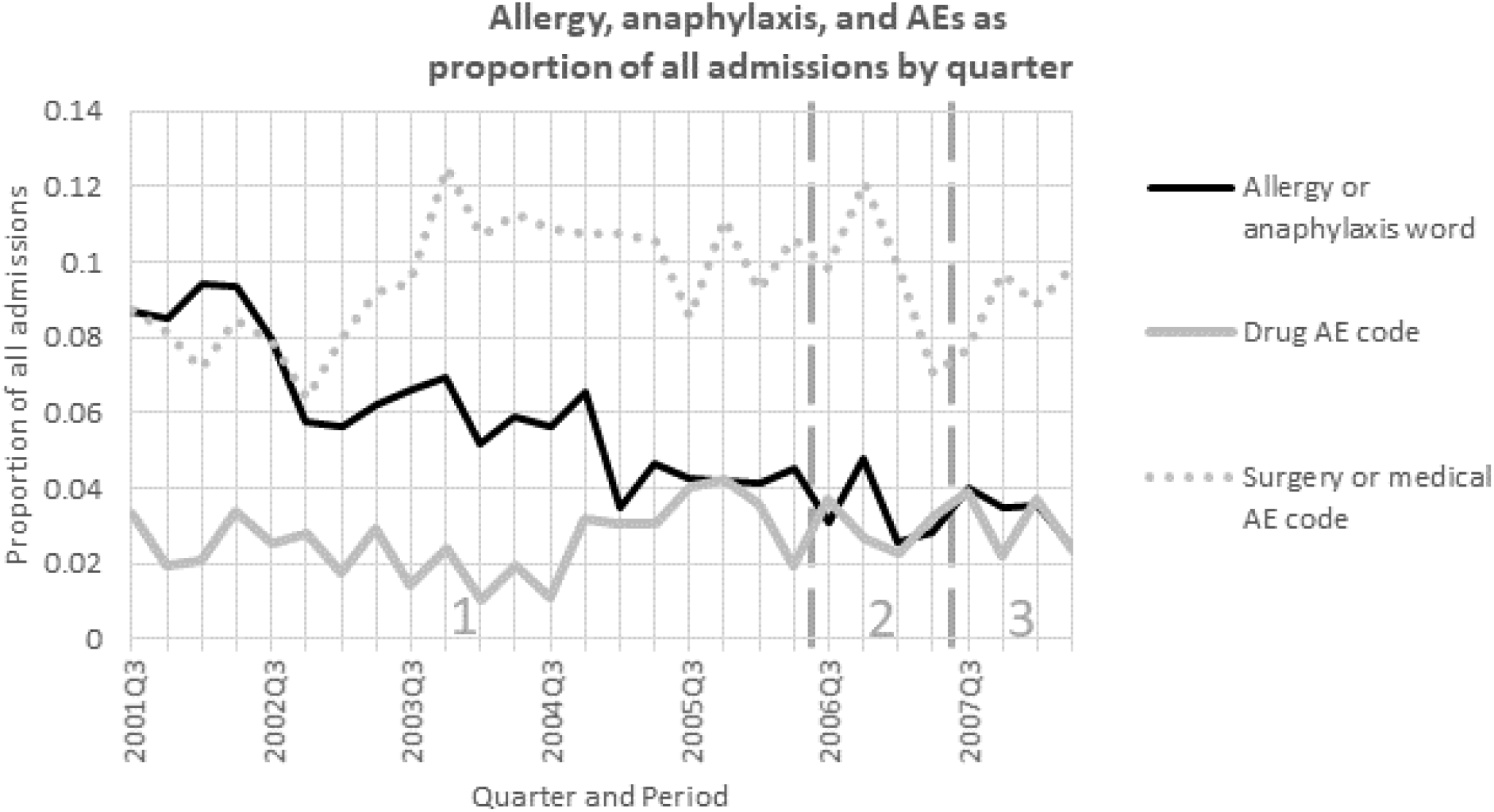
Allergy, anaphylaxis, and AE as proportion of admissions by quarter. For allergy or anaphylaxis word, slope=-0.0022 (95%CI −0.0027 to −0.0018), p<0.0001. For drug AE code, slope=0.00031 (95%CI - 0.000079 to 0.00070), p=0.12. For surgery or medical AE code, slope=0.00049 (95%CI −0.00022 to 0.0012), p=0.18. Figure 7 notes: • Allergy or anaphylaxis word: “allerg*” or “anaphyl*” • Drug AE code: 960** to 979** Poisoning By Drugs, Medicinals And Biological Substances • Surgery or medical AE code: 996** to 999** Complications Of Surgical And Medical Care, Not Elsewhere Classified

The other rare and infrequent terms, related diagnosis or procedure codes, and foreign language sentences were rare throughout all three time periods and increased during period 3.

## DISCUSSION

We succeeded in our expectation of finding increases in clinical events and our hope of finding increases in AEs, especially AEs that would not have been reported because they were not attributed. We found increases in hypotension following heparin or presumed-heparin exposure. Hypotension occurring in the cardiac catheterization lab could be a vasovagal reaction [56]. However, vasovagal reaction generally does not respond to fluids and drugs for raising blood pressure, and all our observed patients’ hypotension did respond to treatment. Hypotension can occur as anaphylaxis begins and, alone, may reflect mild anaphylaxis. We note that the nurses and physicians that described the sequence of events did not link sudden hypotension to heparin and the diagnosis codes did not reflect any awareness of a link. The warnings from FDA and the Centers for Disease Control and Prevention about heparin in the winter of 2007-2008 were for anaphylaxis due to adulterated heparin [57, 58]. Knowledge of the extent of the distribution of adulterated heparin products was not specific, so it may have been in the hospital’s stock at the time. We had expected to see increases starting in 2006 because a few articles indicate heparin may have been adulterated before 2007 [59-61], but were surprised that the increases had started before 2006. The reduction in the last quarter coincided with recalls of contaminated heparin products and lend credibility to the idea that contaminated heparin was in slowly increasing use at this hospital for many years. We are struck that such a high proportion of the invasive cardiac catheter patients in the last two years experienced hypotension following heparin exposure (either as explicitly documented administration or implicitly in the catheter coating).

The types of clinical event changes we detected from period 2 to period 3 were: increases in patients with common conditions (heart disease, brain injuries, trauma, and complex conditions associated with long hospital stays), increases in rare conditions, change in administration (foreign language portion), and adverse events of concern.

The increases in common conditions may have reflected hospital marketing [62].

The increases in rare conditions could have reflected chance, or marketing as a referral center.

Nine of the adverse events happened outside the hospital and illustrate the utility of hospital records for monitoring severe reactions that occur in other health facilities or outside the healthcare system. Our method was useful for detecting words that are rare in hospital records, partly reflecting events that normally occur outside the hospital.

The topic with the highest document score exhibited typical behavior of a topic containing words that are common to most documents. The filter that was removing words comprised of only digits also removed digits from some words. This resulted in some high frequency words getting into the vocabulary. When topic modeling, this resulted in high scores for these common words in the topics where they were correlated (as expected this happens in several topics) and created a common word topic (topic 18). This topic is a noise topic; the LDA model will put words that are low scoring and not correlated with other topics into their own noise topic in order to deal with noise and frequent words. Because this topic included words that were frequent in almost all documents, as expected the document topic scores for this topic were high [63]. This was dealt with by looking at the other more coherent topics that were assigned to each document (essentially ignoring this common-noise topic, capturing what most documents have in common. The top scoring words in this topic that were general survived the ensemble filtering method as an artifact of the digit-removal step. For future work, we recommend removing this step from the filtering process and relying on the classification terms to filter out irrelevant variations of terms.

Our method worked despite:

• the known challenges posed by clinical text notes

• restriction to one major hospital

• lack of all surgical and non-CCU nursing notes, and variable lack of physician, nursing, or discharge summary notes, probably reflecting hospital policy of gradually converting types of notes to EHRs [64]

• errors up to several weeks in dates.

Different, and hopefully improved, results may be derived from EHRs databases that are more complete and have actual dates.

Much of our manual work to evaluate topics could be reduced with a combination of natural language processing and dictionaries of clinical terms. Dictionaries should include standard acronyms and common abbreviations, and should try to account for context when the meaning of term could be ambiguous. The ability to decipher ongoing care notes will be important for noticing unrecognized signals of AEs.

## CONCLUSIONS

We suggest that heparin contamination may have occurred earlier than previously recognized in the winter of 2007-2008, at a lower rate.

Our method successfully aided in the detection of a variety of medical product AEs that were not attributed in clinicians’ notes, suggesting that this method could be a useful supplement to existing post-marketing surveillance programs at local as well as national levels. The method also found other changes in clinical care experiences. The method is easy to execute and understand and could be adopted by subnational public and private entities. It finds potential adverse events that are candidates for causality assessment with epidemiology or other clinical studies.

Our method enabled manual review of key EHRs by narrowing interest from the original large volume of words used in notes. Future improvements could include automation of the manual review process.

## Data Availability

The study is not a clinical trial. The data are available from the Massachusetts Institute of Technology at MIMIC-III Critical Care Database. https://mimic.physionet.org/about/mimic/.

## ACKNOWLEDGEMENTS

We thank enthusiastic support by our FDA and Booz Allen Hamilton supervisors, Department of Health and Human Services innovation programs (Ignite Accelerator and Data Science CoLab), and Alistair Johnson, DPhil, of the MIMIC-III program, Massachusetts Institute of Technology. George Plopper, PhD, of Booz Allen Hamilton, provided project and consultation support. Many FDA colleagues offered ideas and feedback regarding the selection of the case and the final paper. All authors had access to the data. All authors are responsible for the study topic, design, and interpretation. Dr. Bright, Ms. Dowdy, Dr. Rankin, and Dr. Blok are responsible for data processing and analysis.

## Conflict of interest and disclaimer

The research was done with FDA support and under contract HHSF223201510027B between FDA and Booz Allen Hamilton Inc. None of the authors have other relevant financial interests. The opinions are those of the authors and do not represent official policy of either the FDA or Booz Allen Hamilton.

## ABBREVIATIONS USED MORE THAN ONCE

AE: Adverse events
AF: Atrial fibrillation
BIDMC: Beth Israel Deaconess Medical Center
CABG: Coronary artery bypass graft
CCU: Critical (or Intensive) Care Unit
CPR: Cardiopulmonary resuscitation
DMII: Diabetes mellitus, type 2
DVT: Deep vein thrombosis
EHRs: Electronic healthcare records
FDA: Food and Drug Administration
HD: Hospital day
HIT: Heparin induced thrombocytopenia
IABP: Intra-aortic balloon pump
IPH: Intraparenchymal hemorrhage
IV: Intravenous
LDA: Latent Dirichlet Allocation algorithm for topic modeling
LR: Logistic regression supervised learning algorithm
MCA: Middle cerebral artery
MIMIC-III: Medical Information Mart for Intensive Care III
MRI: Magnetic resonance image
MVA: Motor vehicle accident MVC Motor vehicle collision
NB: Naïve Bayes supervised learning algorithm
NLP: Natural language processing
O2: Oxygen
OR: Operating room
PAE: Potential adverse event
PICC: Peripherally inserted central catheter
POD: Post-operative day
tPA: Tissue plasminogen activator
UTI: Urinary tract infection

